# Racial and ethnic disparities for SARS-CoV-2 positivity in the United States: a generalizing pandemic

**DOI:** 10.1101/2021.04.27.21256215

**Authors:** Jacqueline M. Ferguson, Amy C. Justice, Thomas F. Osborne, Hoda S. Abdel Magid, Amanda L. Purnell, Christopher T. Rentsch

## Abstract

The coronavirus pandemic has disproportionally impacted racial and ethnic minority communities in the United States. These disparities may be changing over time as outbreaks occur in different communities. Using electronic health record data from the Department of Veterans Affairs, we estimated odds ratios, stratified by region and time period, for testing positive for SARS-CoV-2 among 951,408 individuals tested for SARS-CoV-2 between February 12, 2020 and February 12, 2021. Our study found racial and ethnic disparities for testing positive were most pronounced at the beginning of the pandemic and decreased over time. A key finding was that the disparity among Hispanic individuals attenuated but remained elevated over the entire study period. We identified variation in racial and ethnic disparities in SARS-CoV-2 positivity by time and region independent of underlying health status and other key factors in a nationwide cohort, which provides important insight for strategies to contain and prevent further outbreaks.

## Introduction

The coronavirus pandemic has disproportionally impacted racial and ethnic minority communities in the United States.^1–3^ Evidence has highlighted the vast disparities in SARS-CoV-2 infection and subsequent COVID-19 among persons who were Black, Hispanic, or Native Hawaiian/Pacific Islander.^4–7^ Recently, additional analyses have suggested that racial and ethnic disparities may be changing over time as outbreaks spread from racially and ethnically diverse metropolitan centers to more rural and less diverse areas.^4,5,8^ In this report, we updated our previous analyses^4,5^ to evaluate changes in disparities for testing positive with SARS-CoV-2 over the first full year of the pandemic and by geographic region in the largest integrated healthcare system in the United States.

## Methods

Using national electronic health record data from the Department of Veterans Affairs (VA), we conducted a retrospective cohort analysis of all individuals tested for SARS-CoV-2 between February 12, 2020 and February 12, 2021, inclusive. Methods have been previously described in detail.^4,5^ In brief, we used multivariable logistic regression to estimate odds ratios (OR) and 95% confidence intervals (CI) for testing positive for SARS-CoV-2 for non-Hispanic Black, Hispanic, Asian, American Indian/Alaska Native. Native Hawaiian/Pacific Islander, and people of mixed race, relative to non-Hispanic White individuals. All models were adjusted for other personal characteristics (sex, age, rural/urban residence) and a wide range of clinical characteristics, including baseline comorbidity (asthma, cancer, chronic kidney disease, chronic obstructive pulmonary disease, diabetes mellitus, hypertension, liver disease, vascular disease), substance use (alcohol consumption, alcohol use disorder, smoking status), medication history (angiotensin converting enzyme inhibitor, angiotensin II receptor blocker). Models were additionally conditioned on VA site of care to account for spatial differences in SARS-CoV-2 burden.

Models were a priori stratified into three waves based on the temporal distribution of SARS-CoV-2 cases nationally: February 12 – May 31, 2020 (wave 1); June 1 – September 30, 2020 (wave 2); October 1, 2020 – Feb 12, 2021 (wave 3). Due to the size of the third national wave, we split this period into two waves containing roughly equal numbers of SARS-CoV-2 cases (October 1 – December 11, 2020 (wave 3a); and December 12, 2020 – February 12, 2021 (wave 3b).

To evaluate regional differences in the most recent wave (3b: December 12, 2020 – February 12, 2021), models were further stratified by US Census region (i.e., West, South, Midwest, and Northeast). Due to low number of events, we combined American Indian/Alaska Native, Native Hawaiian/Pacific Islander, and patients of mixed race into an “other” category for these models. Data analysis was performed using SAS version 9.4 (SAS Institute, Cary, NC).

## Results

Of 951,408 individuals tested during the study period, 111,912 (11.8%) tested positive for SARS-CoV-2 (**Table 1**). All non-White groups had higher crude prevalence of positive tests than White individuals (10.8%), with the largest differences observed among Black (13.0%), Hispanic (15.4%), and American Indian/Alaska Native (13.2%) groups. By region, the crude prevalence of positive tests was highest in Midwest (13.7%) and lowest in Northeast (10.3%). Individuals who were younger or male had a slightly higher crude prevalence of positive tests than those who were older or female. Over time, the prevalence of positive tests increased from 6.9% in wave 1 and 6.2% in wave 2 to 14.8% in wave 3a and 22.0% in wave 3b. In wave 1, all racial and ethnic minority groups had a higher unadjusted percentage of positive tests relative to White individuals, with the highest percentage observed among Black individuals at 11.8% (**Figure 1**). In wave 3b, the percentage of positive tests among White individuals remained lower than all other racial and ethnic minority groups, with the highest percentage observed among Hispanic individuals at 26.3%. Over the study period, White individuals experienced the steepest increase in test positivity percentage (a 4.7-fold relative increase from 4.5% in wave 1 to 21.0% in wave 3b) compared to increases observed in all other racical and ethnic minority groups.

**Table 1.**
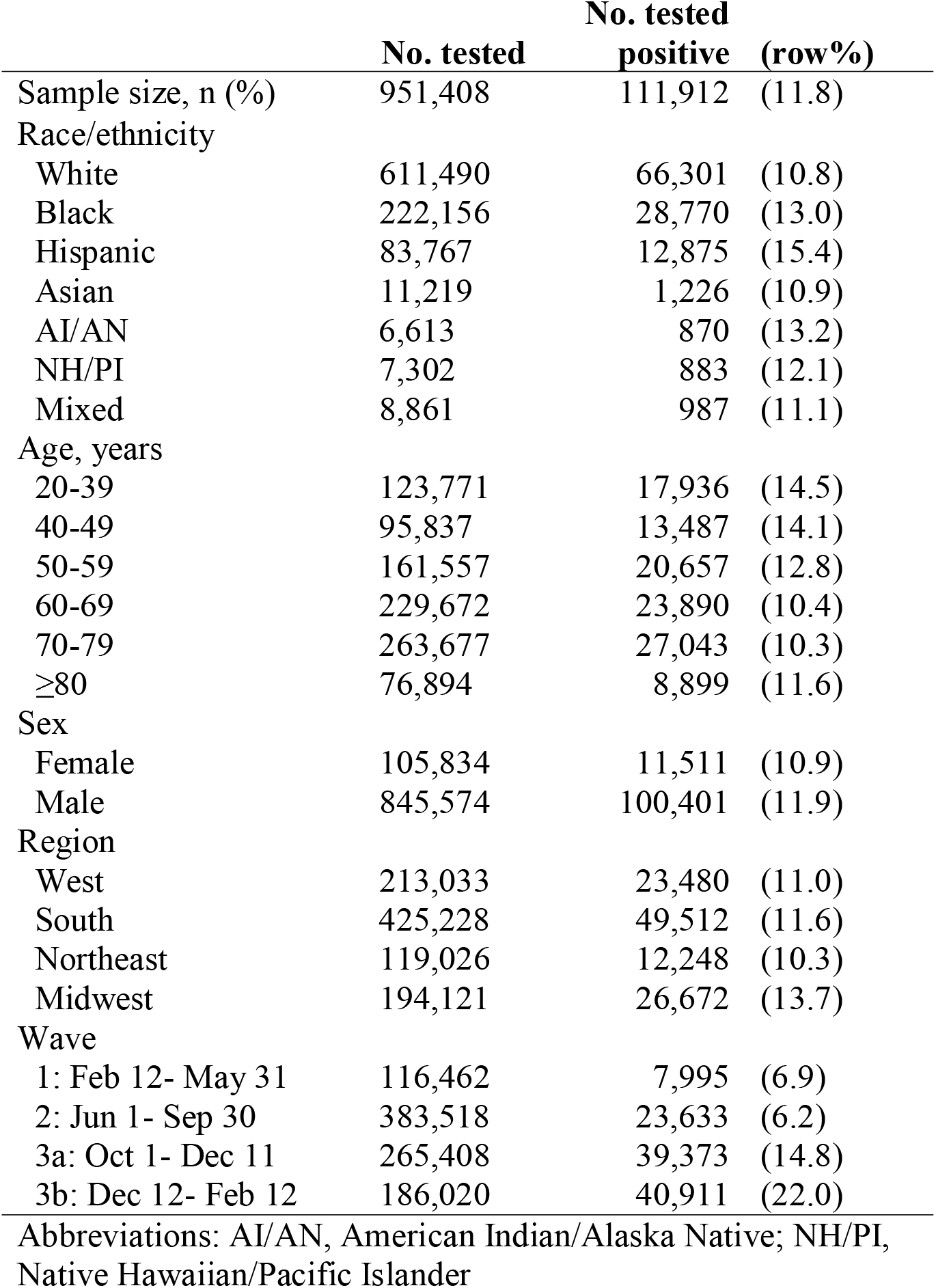
Characteristics of all individuals tested and tested positive for SARS-CoV-2 between February 12, 2020 and February 12, 2021

|  | **No. tested** | **No. tested**<br>**positive** | **(row%)** |
| --- | --- | --- | --- |
| Sample size, n (%) | 951,408 | 111,912 | (11.8) |
| Race/ethnicity |  |  |  |
| White | 611,490 | 66,301 | (10.8) |
| Black | 222,156 | 28,770 | (13.0) |
| Hispanic | 83,767 | 12,875 | (15.4) |
| Asian | 11,219 | 1,226 | (10.9) |
| AI/AN | 6,613 | 870 | (13.2) |
| NH/PI | 7,302 | 883 | (12.1) |
| Mixed | 8,861 | 987 | (11.1) |
| Age, years |  |  |  |
| 20-39 | 123,771 | 17,936 | (14.5) |
| 40-49 | 95,837 | 13,487 | (14.1) |
| 50-59 | 161,557 | 20,657 | (12.8) |
| 60-69 | 229,672 | 23,890 | (10.4) |
| 70-79 | 263,677 | 27,043 | (10.3) |
| ≥80 | 76,894 | 8,899 | (11.6) |
| Sex |  |  |  |
| Female | 105,834 | 11,511 | (10.9) |
| Male | 845,574 | 100,401 | (11.9) |
| Region |  |  |  |
| West | 213,033 | 23,480 | (11.0) |
| South | 425,228 | 49,512 | (11.6) |
| Northeast | 119,026 | 12,248 | (10.3) |
| Midwest | 194,121 | 26,672 | (13.7) |
| Wave |  |  |  |
| 1: Feb 12- May 31 | 116,462 | 7,995 | (6.9) |
| 2: Jun 1- Sep 30 | 383,518 | 23,633 | (6.2) |
| 3a: Oct 1- Dec 11 | 265,408 | 39,373 | (14.8) |
| 3b: Dec 12- Feb 12 | 186,020 | 40,911 | (22.0) |

Abbreviations: AI/AN, American Indian/Alaska Native; NH/PI, Native Hawaiian/Pacific Islander

**Figure 1:**
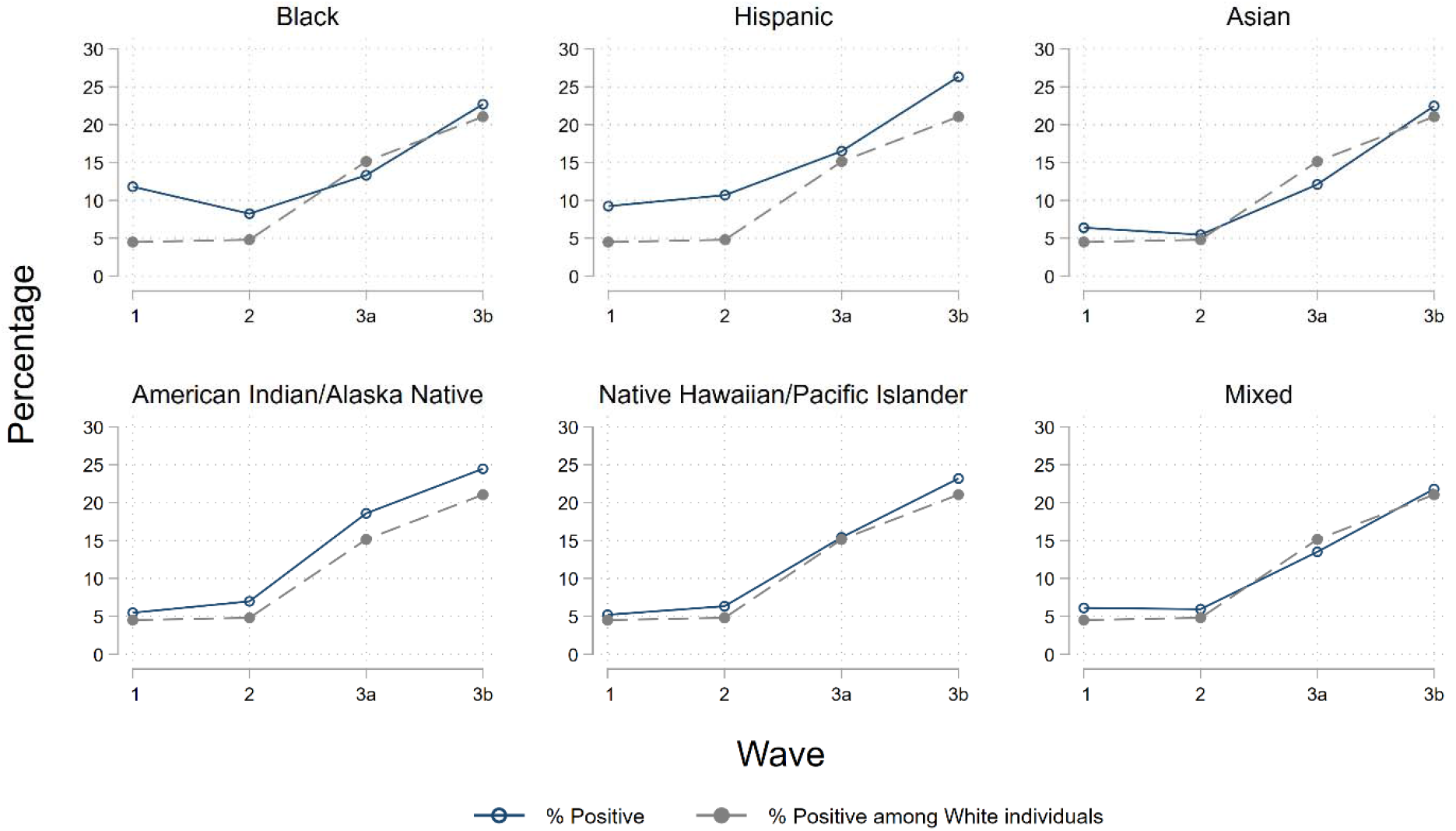
Unadjusted race and ethnicity specific SARS-CoV-2 test positivity percentage by
wave between February 12, 2020 and February 12, 2021 Wave 1 (February 12 – May 31, 2020); Wave 2 (June 1 – September 30, 2020); Wave 3a (October 1 –December 11, 2020); and Wave 3b (December 12, 2020 – February 12, 2021).

After adjustment for personal and clinical characteristics, those who were Black (OR 1.22, 95% CI 1.20-1.25), Hispanic (1.50, 1.47-1.54), American Indian/Alaska Native (1.19, 1.11- 1.28), or Native Hawaiian/Pacific Islander (1.17, 1.08-1.25) had elevated odds of testing positive, with no evidence of disparities among Asian individuals (0.99, 0.93-1.05) or people of mixed race (1.00, 0.93-1.07) compared to White individuals over the entire study period. However, there was substantial variation over time. Disparities for testing positive decreased for all racial and ethnic minorities over the study period (**Figure 2**) with the largest disparities present in wave 1. In wave 1, disparities in test positivity were observed among Black (1.98, 1.86-2.10), Hispanic (1.88, 1.71-2.06), Asian (1.42, 1.11-1.82), and American Indian/Alaska Native (1.72, 1.26-2.34) individuals compared to White individuals. There was some suggestion of a disparity for testing positive among Native Hawaiian/Pacific Islander individuals (1.32, 0.96-1.82) and no observed disparity among people of mixed race (1.15, 0.89-1.50) in wave 1. In wave 3b, disparities for testing positive were not observed among any racial or ethnic minority group, apart from Hispanic individuals (1.34, 1.28-1.40). A notable decrease in test positivity disparity was seen among Black individuals, from a near doubling of odds for testing positive in wave 1 (1.98, 1.86-2.10) to only marginally elevated odds by wave 3b (1.03, 1.00-1.06) compared to White individuals.

**Figure 2:**
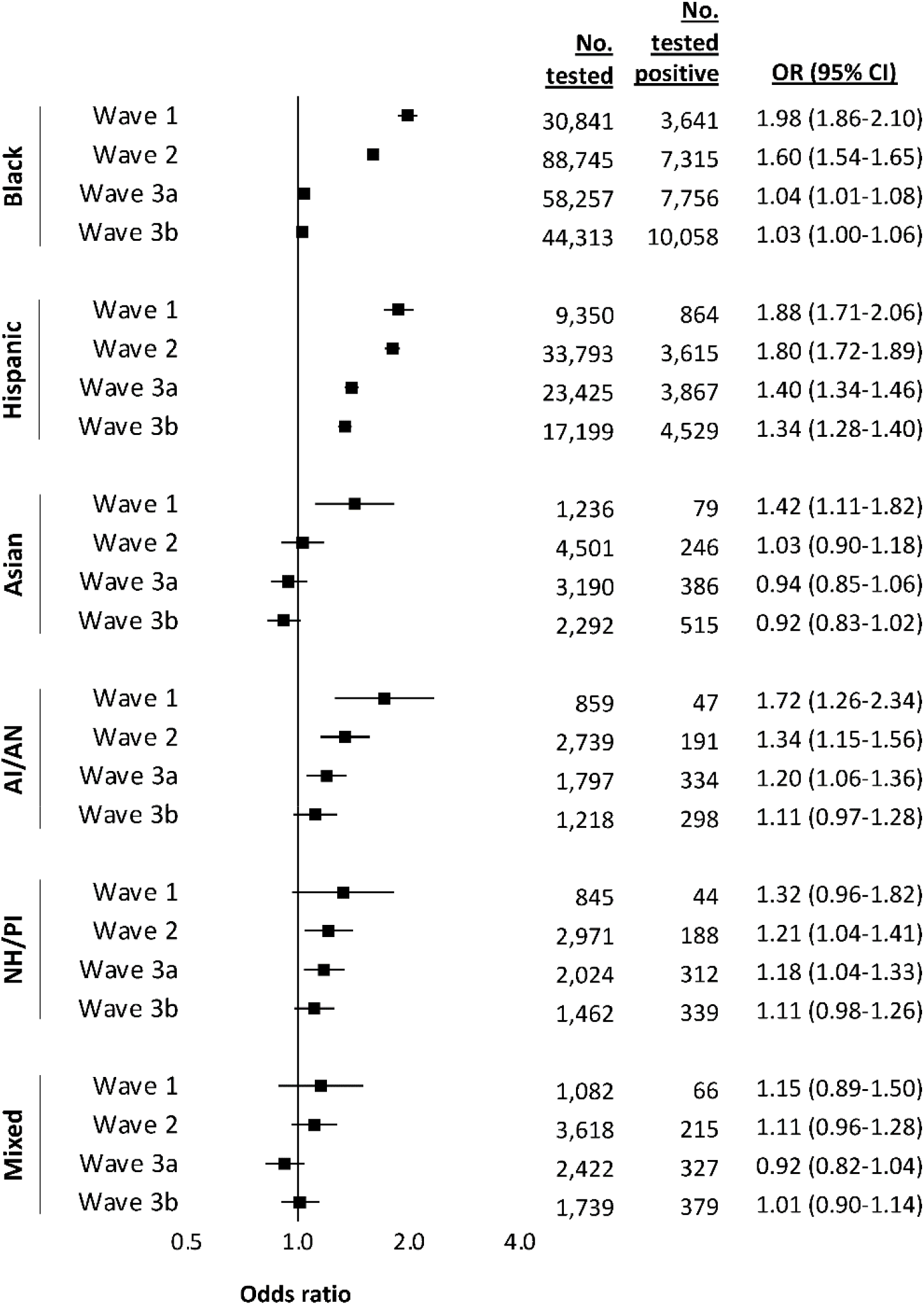
Adjusted racial and ethnic disparities in testing positive for SARS-CoV-2 between February 12, 2020 and February 12, 2021, by wave of the pandemic *Notes*: Wave 1 (February 12 − May 31, 2020); Wave 2 (June 1 − September 30, 2020); Wave 3a (October 1 −December 11, 2020); and Wave 3b (December 12, 2020 − February 12, 2021). Referent group for all comparisons is white. Models conditioned on site of care and adjusted for other demographics, baseline comorbidity, substance use, and medication history. *Abbreviations:* OR, odds ratio; CI, confidence interval; AI/AN, American Indian/Alaska Native; PI/NH, Pacific Islander/Native Hawaiian/Pacific Islander.

We found some evidence of regional variation in the disparity for testing positive in the most recent wave (**Figure 3**). Black individuals and people of other race (i.e., American Indian/Alaska Native, Native Hawaiian/Pacific Islander, and mixed race) had marginally higher odds for testing positive in the South (1.07, 1.03-1.11 for Black; 1.13, 1.01-1.26 for other race). Disparities for testing positive among Hispanic individuals were present in all regions, most notably in the West (1.49, 1.39-1.59). There was no evidence of variation in disparities for testing positive across geographic regions for Asian individuals; however, confidence intervals were wide due to low numbers testing positive for this group in most regions.

**Figure 3:**
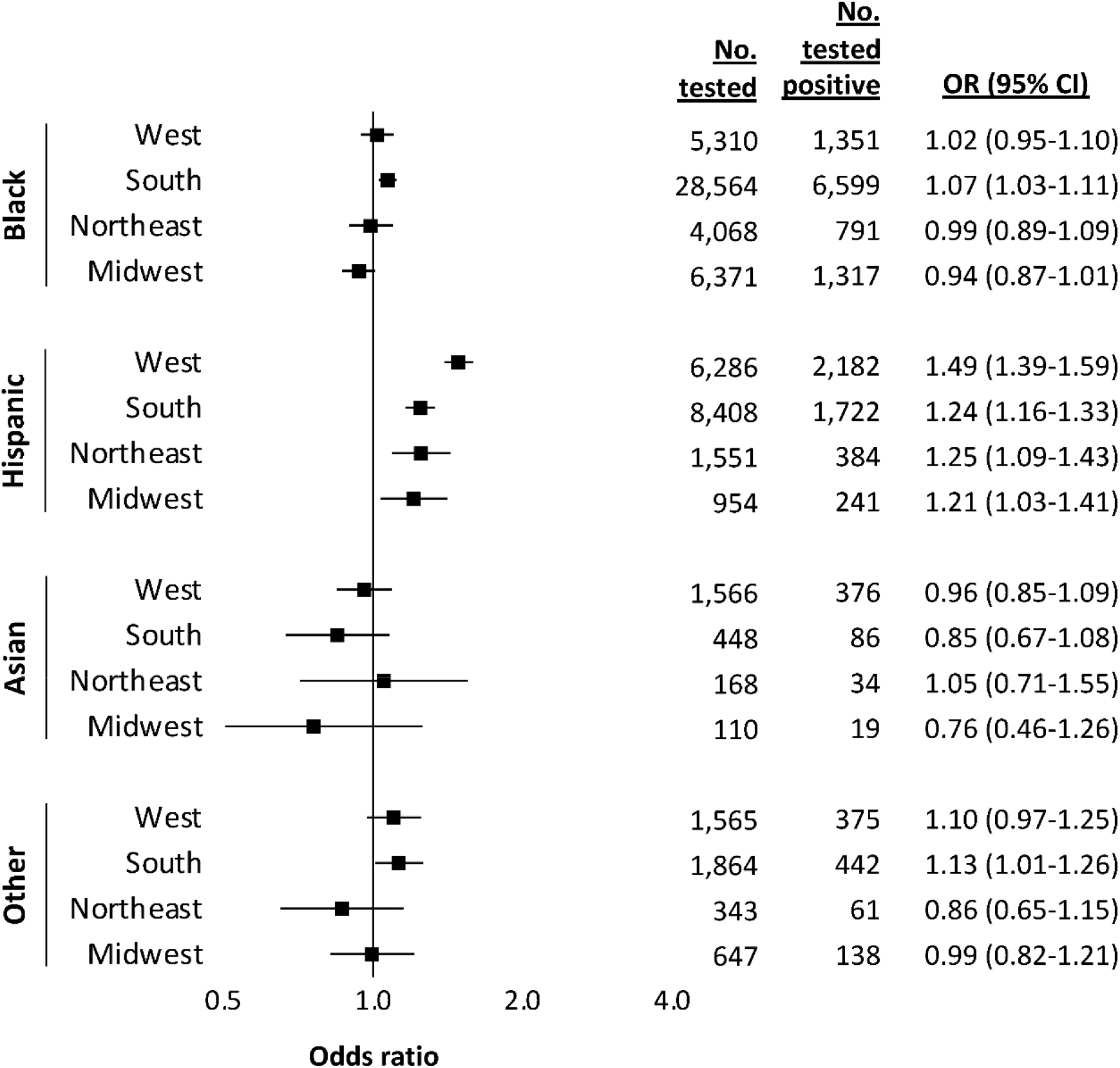
Adjusted racial and ethnic disparities in testing positive for SARS-CoV-2 between December 12, 2020 and February 12, 2021, by region *Notes:* Referent group for all comparisons is White. “Other” category included American Indian/ Alaska Native, Pacific Islander/Native Hawaiian. And Mixed. Models conditioned or of care and adjusted for other demographics, baseline comorbidity, substance use, and medication history. *Abbreviations:* OR, odds ratio; CI, confidence interval.

## Discussion

Our study found that racial and ethnic disparities for testing positive for SARS-CoV-2 were most pronounced at the beginning of the pandemic and that these disparities decreased over time after accounting for a wide range of personal and clinical characteristics. By the end of the first 12 months of the pandemic, disparities for testing positive were attenuated but remained elevated for Hispanic individuals and were no longer observed for any other group. This attenuation in disparities appears to be due, at least in part, to a stronger relative increase in the test positivity percentage among White individuals over time rather than a decline in test positivity among racial and ethnic minority groups, which is likely driven by the pandemic generalizing from diverse metropolitan areas to less diverse rural areas.^4,^^5,^^8^

Our findings on disparities for testing positive among Black and Hispanic individuals in the first months of the pandemic have been demonstrated previously.^1–5^ This study extended previously published models to evaluate patterns in disparities over the first full year of the pandemic. A novel finding was that disparities for testing positive dramatically attenuated and were no longer observed among all racial and ethnic groups apart from Hispanic individuals. Another novel finding was the identification of disparity among Asian individuals in the first wave of the pandemic, which was obscured in the time-pooled model.

A key finding was that the disparity for testing positive among Hispanic individuals attenuated but remained elevated over the study period. In the last wave assessed between December 2020 and February 2021, Hispanic individuals had 34% elevated odds for testing positive relative to White individuals. This persistent disparity among Hispanic individuals was observed across all geographic regions, with the greatest disparity in the West. A deeper understanding of the mechanism for this association is needed but may be due to the lack of nationwide media coverage and targeted and appropriate outreach to Hispanic populations in the United States. Hispanic individuals are also overrepresented in essential and frontline jobs, which increases their likelihood of SARS-CoV-2 exposure and they may face barriers (e.g., precarious employment or financial limitations) to taking sick leave that would help reduce the spread of SARS-CoV-2.^9^

Our findings of racial and ethnic disparities for testing positive for SARS-CoV-2 provide important insight to help tailor strategies to contain and prevent further outbreaks in the United States. Early in the pandemic, tailored interventions to groups with higher risks may have been most effective. Now that the epidemic has generalized from large metropolitan centers with very high incidence to a more consistent rate of incidence across the country, racial and ethnic groups may be affected more equally suggesting that widescale prevention interventions for all persons may be most effective. However, the continued disparities among Hispanic groups suggest that targeted assessment and data informed interventions are required. Furthermore, while there is a more consistent rate of SARS-CoV-2 across the United States, targeted assessment may still be useful for curtailing local infection hotspots. SARS-CoV-2 is impacting all communities and is now much less concentrated in specific vulnerable groups compared to early in the pandemic. This does not imply that the overall cumulative burden of COVID-19 may be equal, as marginalized populations such as persons of color experienced substantial excess rates earlier in the epidemic and may experience excess extended effects from infection. Of note, access to free or subsidized care at VA may help reduce the impact of negative social determinants of health as prior reports found no racial or ethnic disparities in mortality among patients who tested positive for SARS-CoV-2 in the VA.^4^

The VA electronic health record database offers the single largest nationwide data resource available in the United States with the necessary information on system-wide testing and detailed medical histories to examine racial and ethnic disparities. Our analysis identified time and regional variation in racial and ethnic disparities in SARS-CoV-2 positivity over the first full year of the pandemic independent of underlying health status and other key factors in a large, nationwide cohort. However, our analysis should be interpreted with some limitations, including those we previously described in detail.^4,5^ In brief, we only examined tests administered in the VA; therefore, our results may not be representative of all Veterans tested for SARS-CoV-2. Second, although this population was primarily male, it included over 100,000 women. Third, as is the case with most electronic health record data sources, we did not have the necessary information to account for social determinants of health (e.g., occupation or household details) in our analysis, which are critical to understanding and preventing health inequities, particularly in infectious disease outbreaks. Careful research is needed to evaluate the association between social determinants of health and disparities seen during the COVID-19 pandemic as they may operate as confounders or may be on the causal pathway.

## Data Availability

Due to US Department of Veterans Affairs (VA) regulations and our ethics agreements, the analytic data sets used for this study are not permitted to leave the VA firewall without a Data Use Agreement. This limitation is consistent with other studies based on VA data. However, VA data are made freely available to researchers with an approved VA study protocol. For more information, please visit https://www.virec.research.va.gov or contact the VA Information Resource Center at.

## Author Contributions

**Concept and Design:** JMF, CTR, ACJ, TFO

**Drafting of the manuscript:** JMF, CTR

**Acquisition, analysis, or interpretation of the data:** CTR, ACJ, JMF, TFO

**Critical revision of the manuscript for important intellectual content:** All authors

**Statistical Analysis:** CTR, JMF

**Administrative, technical, or material support:** ACJ, TFO

**Supervision:** ACJ, TFO

## Disclaimer

Views expressed are those of the authors and the contents of this article do not represent the views of the US Department of Veterans Affairs or the United States Government.

## Ethics

This study was approved by the institutional review boards of VA Connecticut Healthcare System (VA AJ0013) and Yale University (1506016006). It has been granted a waiver of informed consent and is Health Insurance Portability and Accountability Act compliant.

